# Interrogating Changes in COVID-19 Vaccine Intent During the National Vaccine Rollout in the Philippines

**DOI:** 10.1101/2024.01.20.24301539

**Authors:** Alexandria Caple, Arnie Dimaano, Marc Martin Sagolili, April Anne Uy, Panjee Mariel Aguirre, Dean Alano, Kaila Andrine Baul, Giselle Sophia Camaya, Angelo Raine Carlos, Brent John Ciriaco, Princess Jerah Mae Clavo, Dominic Cuyugan, Cleinne Florence Geeseler Fermo, Paul Jeremy Lanete, Ardwayne Jurel La Torre, Thomas Loteyro, Raisa Mikaela Lua, Nicole Gayle Manansala, Raphael Mosquito, Alexa Marie Octaviano, Alexandra Erika Orfanel, Anne Elizabeth Papa, Gheyanna Merly Pascual, Mark Nhel Peralta, Aubrey Joy Sale, Sophia Lorraine Tendenilla, Maria Sofia Lauren Trinidad, Nicole Jan Trinidad, Daphne Louise Verano, Nicanor Austriaco

**Author notes:** Corresponding Author: Rev. Nicanor Austriaco, OP, UST Laboratories for Vaccine Science, Molecular Biology, and Biotechnology, University of Santo Tomas, España Blvd., Manila 1015, PHILIPPINES.

## Abstract

To mitigate the unprecedented health, social, and economic damage of COVID-19, the Philippines implemented a nation-wide vaccine program to mitigate the effects of the global pandemic. In a previous study, we interrogated COVID-19 vaccine intent in the country by deploying a nationwide open-access online survey, two months before the initial rollout of the national vaccination program that began in March of 2021. In this follow-up study, we explored the influence of the ongoing vaccine rollout on vaccine intent by deploying a similar survey six months after the initial rollout of vaccines had begun throughout the archipelago. Our data suggests that the presence of vaccines and vaccinated individuals in a Filipino community predicts vaccination intent. When directly asked if they were more willing to receive a vaccination than six months prior, 92.26% agreed to some extent that they were indeed more willing. Finally, despite the changes in the numbers of respondents who were more open to the COVID-19 vaccines, there were no significant changes in the predictive power of our Health Belief Model (HBM) constructs. For the most part, the HBM factors that predicted vaccination intent in our earlier study also predicted vaccination intent in this current study.

## Introduction

On January 30, 2020, the Department of Health (DOH) of the Philippines reported its first case of COVID-19, a novel respiratory disease first identified in Wuhan, China, that is caused by the coronavirus, SARS-CoV-2 (Xie et al., 2020; Zhu et al., 2020; Guan et al., 2020; Reis et al., 2022). With widespread human-to-human transmission, the virus is highly contagious, and the COVID-19 pandemic became a global concern of historic proportions (van Ginneken et al., 2022; Siddiqui, Alhamdi & Alghamdi, 2022; Sachs et al., 2022; Cassell et al., 2022). As of April 5, 2023, there have been 4,082,418 confirmed cases and 66,390 confirmed deaths from COVID-19 reported by the DOH throughout the archipelago (www.covid19.gov.ph).

Vaccination has long been regarded as the most effective means for combating infectious disease (Rappuoli et al., 2014; Sathyanarayana et al., 2020). The Philippines began its national vaccine drive against COVID-19 on March 1, 2021, with the goal of vaccinating seventy million of its citizens by the end of the calendar year (Inter-Agency Task Force for the Management of Emerging Infectious Disease, 2021). One of the ongoing challenges for this campaign is the vaccine hesitancy among the Filipino people (Alfonso et al., 2021). Though immunization rates had been relatively high in the Philippines for many decades, the controversial 2016 rollout of the dengue vaccine, Dengvaxia, triggered significant drops in the rates of immunization as Filipino parents refused to have their children routinely vaccinated against polio, chicken pox, and tetanus (Fatima & Syed, 2018; Smith, 2018; Reñosa et al., 2022).

The Health Belief Model (HBM) posits that people are likely to adopt disease prevention behaviors and to accept medical interventions like vaccines if there is sufficient motivation and cues to action (Rosenstock, Strecher & Becker, 1988; Limbu, Gautam & Pham, 2022a; Limbu & Gautam, 2023). Motivational factors include perceived susceptibility to and severity of the disease and perceived benefits of the vaccine. Cues to action include information, people, and events that nudge the individual towards vaccination. The HBM has been adopted as a conceptual framework that has been used to evaluate the beliefs and attitudes toward a diversity of vaccines including the influenza, human papillomavirus, and hepatitis B vaccines (Teitler-Regev, Shahrabani & Benzion, 2011; Donadiki et al., 2014; Hu et al., 2017; Chen et al., 2019). Moreover, several studies have shown that the HBM constructs can serve as an important predictor of influenza vaccination uptake (Brewer et al., 2007; Shahrabani, Benzion & Yom Din, 2009; Shahrabani & Benzion, 2010; Tsutsui, Benzion & Shahrabani, 2012; Alsuwailem et al., 2023). During the COVID-19 pandemic, the HBM was used to assess the root causes of COVID-19 vaccine hesitancy in the Asia-Pacific region and beyond (Kabir et al., 2021; Alobaidi, 2021; Huynh et al., 2021; Zilhadia et al., 2022; Jian Ng et al., 2022; Shah et al., 2022; Seangpraw et al., 2022b,a; Limbu, Gautam & Pham, 2022b; Ghazy et al., 2022; Hidayana et al., 2022; Qin et al., 2022; Harutyunyan et al., 2023; Berger et al., 2023; Toledo-López et al., 2023).

In an earlier study, we interrogated COVID-19 vaccine intent in the Philippines by deploying a nationwide open-access online survey, two months *before* the rollout of the national vaccination program (Caple et al., 2022). Multivariable analysis of our data revealed then that HBM constructs were associated with vaccination intent in the Filipino population. In this follow-up study, we explored the influence of an ongoing vaccine rollout on vaccine intent by deploying a survey six months *after* the initial rollout of vaccines had begun throughout the Philippines. We wanted to determine if and how a vaccine rollout would predict public perceptions of the vaccines.

Results of the current study suggest that the presence of vaccines and vaccinated individuals in a Filipino community was associated with an increase in vaccination intent. When directly asked if individuals were more willing to receive a vaccination than six months prior, 92.26% agreed to some extent that they were indeed more willing. Finally, despite the changes in the numbers of respondents who were more open to the COVID-19 vaccines, there were no significant changes in the predictive power of the HBM constructs. For the most part, HBM factors that predicted vaccination intent in our earlier study also predicted vaccination intent in this current study.

## Materials & Methods

### Participants and Survey Design

The current study design was a cross-sectional, open-access, anonymous, web-based survey— developed using Qualtrics—conducted from August 14, 2021, to September 13, 2021. Our research team deployed an anonymous survey link via the social platforms of the University of Santo Tomas (UST) like Twitter and Facebook and university mailing lists including the UST School of Science and UST Student Council to distribute the survey. Participants were encouraged to distribute the survey link to their contacts throughout the country. The questionnaire was written in both Filipino and English. Responses used for data collection were limited to respondents who were at least 18 years old. An IRB-approved informed consent statement was included in the survey instrument to welcome respondents who had clicked on the anonymous survey link.

### Survey Instrument

The survey consisted of questions and statements that assessed the following: (1) demographics, health status, and COVID-19 experience; (2) openness to a COVID-19 vaccine, and if participants received a COVID-19 vaccine; (3) perceived susceptibility to and severity of COVID-19; (4) perceived benefits of a COVID-19 vaccine; and (5) barriers of a COVID-19 vaccine, and cues to action to obtain a COVID-19 vaccine.

Demographics, health status, and COVID-19 experience: Demographic information including age, sex, marital status, education level, occupation, and urban or rural location of residence were collected. Participants were also asked if they have an existing chronic condition, how they would rate their overall health, if they ever personally tested positive for COVID-19, to indicate if they know anyone who has tested positive for COVID-19, and if they know someone who has been vaccinated against COVID-19.

Openness to a COVID-19 vaccine and participant vaccination status: Openness to a COVID-19 vaccine was assessed using two items (“Would you take a COVID-19 vaccine?”, “Are you more willing to take a COVID-19 vaccine now than you were six months ago?”) on a five-point scale ranging from 1 = ‘definitely no’ to 5 = ‘definitely yes’. Responses were further recoded into two distinct categories: vaccine resistance (responses included: ‘definitely no’, ‘probably no’, and ‘unsure’) and vaccine acceptance (responses included: ‘probably yes’ and ‘definitely yes’) for both scale items. Participant COVID-19 vaccination status was assessed using a one-item question (“Have you been vaccinated against COVID-19?”). Responses were additionally recoded into three distinct categories: not vaccinated, partially vaccinated, and fully vaccinated. Participants who indicated they had received either partial or full vaccination status were also asked to specify which vaccine they receive (“If you have been vaccinated against COVID-19, which vaccine did you receive?”).

Perceived susceptibility to and severity of COVID-19: Health Belief Model (HBM) derived items were used to assess individual beliefs about a COVID-19 vaccine as well as individual beliefs about a COVID-19 vaccine prior to the national vaccine rollout. Questions posted to participants assessed perceived susceptibility of COVID-19 (three items), and perceived severity of COVID-19 (four items). All response items were on a four-point scale ranging from ‘strongly disagree’ to ‘strongly agree’. For analysis purposes, all responses were coded as either *disagree* (responses included: ‘strongly disagree’ and ‘disagree’) or *agree* (responses included: ‘strongly agree’ and ‘agree’).

Perceived benefits of a COVID-19 vaccine: Perceived benefits were assessed using seven items, including statements that examined perceived benefits of a COVID-19 vaccine compared to before the national vaccine rollout. All response items were rated on a four-point scale ranging from ‘strongly disagree’ to ‘strongly agree’. Similar to perceived susceptibility to and severity of COVID-19, all responses were coded as either *disagree* or *agree*.

Perceived barriers of a COVID-19 and cues to action: Perceived barriers surrounding a COVID-19 vaccine (e.g., “I worry about the possible side-effects of the COVID-19 vaccine”; “I worry about fake COVID-19 vaccines”). For analysis purposes, all responses were coded as either *disagree* or *agree*. Additionally, cues to actions were assessed used eight items. All responses items were on a four-point scale ranging from ‘strongly disagree’ to ‘strongly agree’. All responses were coded as either *disagree* (responses included: ‘strongly disagree’ and ‘disagree’) or *agree* (responses included: ‘strongly agree’ and ‘agree’).

### Ethics Review & IRB Approval

Our study protocol (Protocol Number 21-026) was reviewed and approved by the Institutional Review Board of Providence College on January 15, 2021, with an amendment for the current study being reviewed and approved on July 15, 2021. Originally, we had sought ethical review at the University of Santo Tomas in the Philippines for our initial study but were advised by university authorities there to seek accelerated IRB approval in the United States due to the impact of the then ongoing-COVID-19 pandemic.

### Statistical Analyses

All statistical analyses were conducted using Statistical Package for the Social Sciences (SPSS) version 28. A *p*-value of less than .05 was considered statistically significant. Frequency tables, charts, and proportions were used for data summarization—proportions and their respective 95% confidence intervals (CI) were calculated for each predictor variable. Participant responses to the one-item COVID-19 vaccine status question (“Have you been vaccinated against COVID-19?”) was coded into two groups: not vaccinated and partially vaccinated/fully vaccinated responses. We ran univariate analyses followed by a multinomial logistic regression analysis, including all factors showing significance (*p* < .05), to determine which factors predicted individual COVID-19 vaccine intention. Only significant factors in the univariate analyses were included in the multinomial logistic regression analysis.

## Results

### Demographics

A total of 5,130 complete survey responses were received. Responses received represented participants with diverse demographics (Table 1) and from various regions of the Philippines (Figure 1). Demographic breakdowns were similar to our study of COVID-19 intent three months prior to the national vaccine rollout (Caple et al., 2022). However, the current study sample had a higher representation of younger adults aged 18 to 30 years old (67.3%). Additionally, the majority of participants identified as female (64.2%), single (75.7%), had obtained a college/university degree or above (77.2%), and lived in an urban area (79.7%). A small portion of our sample reported they have an existing chronic condition (12.4%), while 11.9% of respondents reported having ‘fair’ health. Additionally, 64.3% of the sample reported knowing someone who has tested positive for COVID-19.

|  | Have you been vaccinated against COVID-19? |  |  |  |
| --- | --- | --- | --- | --- |
|  | Overall N (%) | Not Vaccinated<br>n = 856<br>(16.7%) | Partially<br>Vaccinated<br>n = 1476<br>(28.8%) | Fully Vaccinated<br>n = 2798<br>(54.5%) |
| *Demographics* |  |  |  |  |
| Age group (years) |  |  |  |  |
| 18–30 | 3453 (67.31) | 701 (20.3) | 1223 (35.4) | 1529 (44.3) |
| 31–40 | 521 (10.16) | 41 (7.9) | 122 (23.4) | 358 (68.7) |
| 41–50 | 451 (8.79) | 52 (11.5) | 65 (14.4) | 334 (74.1) |
| 51–99 | 705 (13.74) | 62 (8.8) | 66 (9.4) | 577 (81.8) |
| Gender |  |  |  |  |
| Female | 3293 (64.19) | 616 (18.7) | 890 (27) | 1787 (54.3) |
| Male | 1837 (35.81) | 240 (13.1) | 586 (31.9) | 1011 (55) |
| Marital Status |  |  |  |  |
| Single | 3883 (75.69) | 741 (19.1) | 1292 (33.3) | 1850 (47.6) |
| Married | 1247 (24.31) | 115 (9.2) | 184 (14.8) | 948 (76) |
| Highest Education Level |  |  |  |  |
| Elementary school or below | 3 (.06) | 1 (33.3) | 2 (66.7) |  |
| Junior high school | 132 (2.57) | 14 (10.6) | 55 (41.7) | 63 (47.7) |
| Senior high school | 1036 (20.19) | 220 (21.2) | 412 (39.8) | 404 (39) |
| College/university of above | 3959 (77.17) | 621 (15.7) | 1007 (25.4) | 2331 (58.9) |
| Occupation |  |  |  |  |
| Blue collar worker | 878 (17.12) | 64 (7.3) | 397 (45.2) | 417 (47.5) |
| Profession/white collar worker | 1627 (31.72) | 133 (8.2) | 328 (20.2) | 1166 (71.7) |
| Self-employed | 541 (10.55) | 62 (11.5) | 209 (38.6) | 270 (49.9) |
| Student | 1710 (33.33) | 548 (32) | 465 (27.2) | 697 (40.8) |
| Housewife/retired/unemployed/other | 374 (7.29) | 49 (13.1) | 77 (20.6) | 248 (66.3) |
| Location |  |  |  |  |
| Rural | 1043 (20.33) | 257 (24.6) | 334 (32) | 452 (43.3) |
| Urban | 4087 (79.67) | 599 (14.7) | 1142 (27.9) | 2346 (57.4) |
| *Experience with COVID-19* |  |  |  |  |
| Ever tested positive for COVID-19 |  |  |  |  |
| Yes | 334 (6.51) | 48 (14.4) | 72 (21.6) | 214 (64.1) |
| No | 4796 (93.49) | 808 (16.8) | 1404 (29.3) | 2584 (53.9) |
| Know anyone who has tested positive for COVID-19 |  |  |  |  |
| Yes | 3299 (64.31) | 617 (18.7) | 649 (19.7) | 2033 (61.6) |
| No | 1831 (35.69) | 239 (13.1) | 827 (45.2) | 765 (41.8) |
| *Health Characteristics* |  |  |  |  |
| Have an existing chronic condition |  |  |  |  |
| Yes | 635 (12.38) | 81 (12.8) | 61 (9.6) | 493 (77.6) |
| No | 4495 (87.62) | 775 (17.2) | 1415 (31.5) | 2305 (51.3) |
| **Perceived overall health** |  |  |  |  |
| Very good | 2021 (39.4) | 268 (13.3) | 703 (34.8) | 1050 (52) |
| Good | 2466 (48.07) | 436 (17.7) | 650 (26.4) | 1380 (56) |
| Fair/Poor/Very poor | 643 (12.53) | 152 (23.6) | 123 (19.1) | 368 (57.2) |

|  | Have you been vaccinated against COVID-19? |  |  |
| --- | --- | --- | --- |
|  | Overall N (%) | Not Vaccinated<br>n = 856<br>(16.7%) | Partially Vaccinated & Fully Vaccinated<br>n = 4274 (83.3%) |
| *Demographics* |  |  |  |
| Age group (years) |  |  |  |
| 18–30 | 3453 (67.31) | 701 (20.3) | 2752 (79.7) |
| 31–40 | 521 (10.16) | 41 (7.9) | 480 (92.1) |
| 41–50 | 451 (8.79) | 52 (11.5) | 399 (88.5) |
| 51–99 | 705 (13.74) | 62 (8.8) | 643 (91.2) |
| Gender |  |  |  |
| Female | 3293 (64.19) | 616 (18.7) | 2677 (81.3) |
| Male | 1837 (35.81) | 240 (13.1) | 1597 (86.9) |
| Marital Status |  |  |  |
| Single | 3883 (75.69) | 741 (19.1) | 3142 (80.9) |
| Married | 1247 (24.31) | 115 (9.2) | 1132 (90.8) |
| Highest Education Level |  |  |  |
| Elementary school or below | 3 (.06) | 1 (33.3) | 2 (66.7) |
| Junior high school | 132 (2.57) | 14 (10.6) | 118 (89.4) |
| Senior high school | 1036 (20.19) | 220 (21.2) | 816 (78.8) |
| College/university of above | 3959 (77.17) | 621 (15.7) | 3338 (84.3) |
| Occupation |  |  |  |
| Blue collar worker | 878 (17.12) | 64 (7.3) | 814 (92.7) |
| Profession/white collar worker | 1627 (31.72) | 133 (8.2) | 1494 (91.8) |
| Self-employed | 541 (10.55) | 62 (11.5) | 479 (88.5) |
| Student | 1710 (33.33) | 548 (32) | 1162 (68) |
| Housewife/retired/unemployed/other | 374 (7.29) | 49 (13.1) | 325 (86.9) |
| Location |  |  |  |
| Rural | 1043 (20.33) | 257 (24.6) | 786 (75.4) |
| Urban | 4087 (79.67) | 599 (14.7) | 3488 (85.3) |
| *Experience with COVID-19* |  |  |  |
| Ever tested positive for COVID-19 |  |  |  |
| Yes | 334 (6.51) | 48 (14.4) | 286 (85.6) |
| No | 4796 (93.49) | 808 (16.8) | 3988 (83.2) |
| Know anyone who has tested positive for COVID-19 |  |  |  |
| Yes | 3299 (64.31) | 617 (18.7) | 2682 (81.3) |
| No | 1831 (35.69) | 239 (13.1) | 1592 (86.9) |
| *Health Characteristics* |  |  |  |
| Have an existing chronic condition |  |  |  |
| Yes | 635 (12.38) | 81 (12.8) | 554 (87.2) |
| No | 4495 (87.62) | 775 (17.2) | 3720 (82.8) |
| **Perceived overall health** |  |  |  |
| Very good | 2021 (39.4) | 268 (13.3) | 1753 (86.7) |
| Good | 2466 (48.07) | 436 (17.7) | 2030 (82.3) |
| Fair/Poor/Very poor | 643 (12.53) | 152 (23.6) | 491 (76.4) |

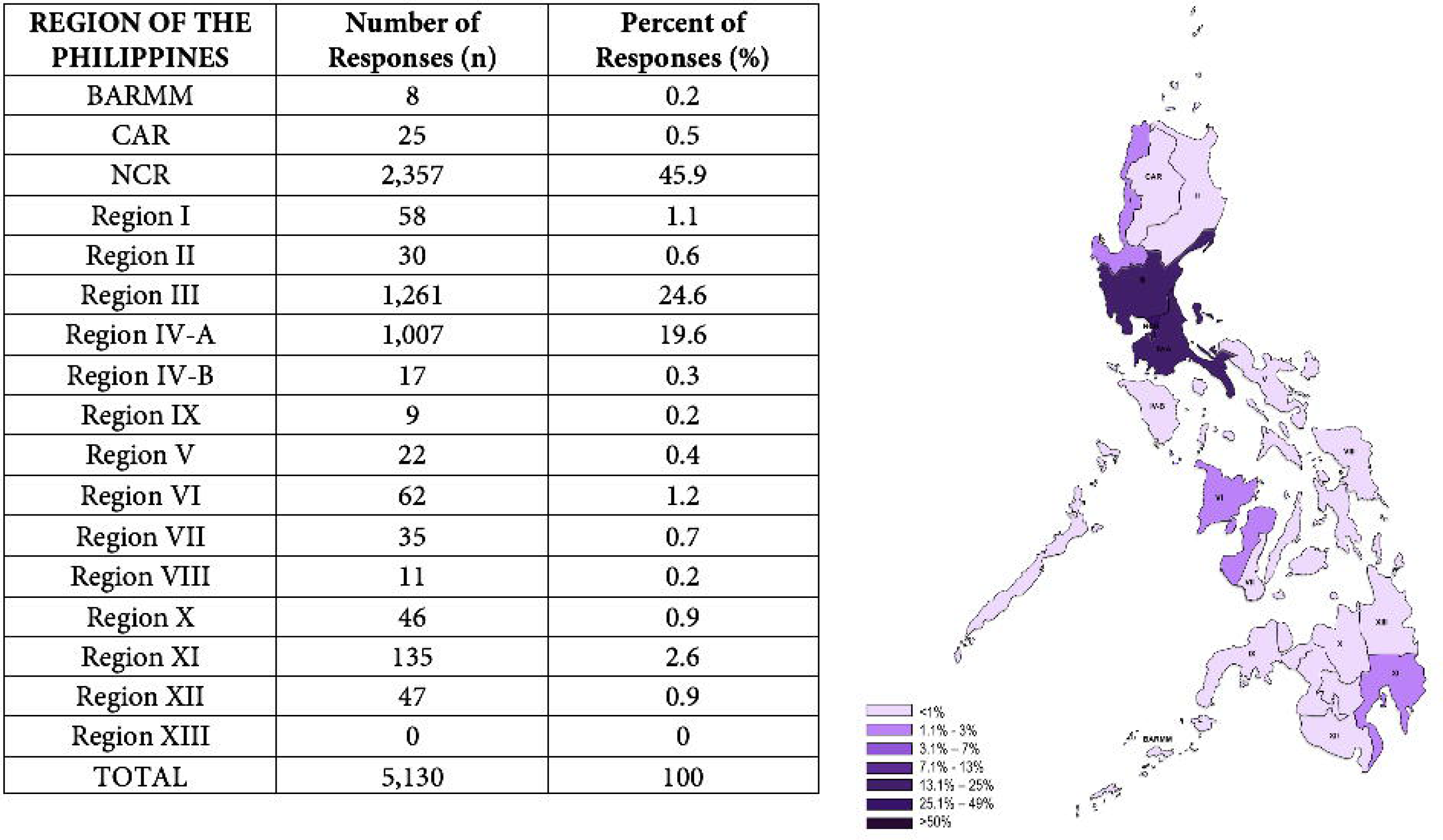

### Health Beliefs

As displayed in Table 2, with regards to perceived susceptibility to COVID-19 vaccines, a portion of respondents (42.8%) agreed to some extent (which includes *strongly agreed/agreed*) there was a high chance of personally contracting COVID-19 in the next few months, which was not significantly different from the 31.5% reported previously (Caple et al., 2022). A significant majority of respondents (81.1%) also agreed to some extent that they were worried about the likelihood of personally contracting COVID-19. This was comparable to the number (84.1%) reported nine months earlier (Caple et al., 2022). Similarly, a large majority of respondents reported agreement to some extent when asked about their personal worry about the likelihood of someone in their family contracting COVID-19 (86.6%). A majority of participants reported COVID-19 is a serious disease with life-threatening complications (95.91%). A notable majority of respondents (92.7%) agreed to some extent that they were personally afraid of contracting COVID-19 and that they would personally be very sick if they contracted COVID-19 (72.1%). These were indistinguishable from the numbers, 93.1% and 75%, that reported previously from our survey completed nine months prior to this study (Caple et al., 2022).

**TABLE 2:**
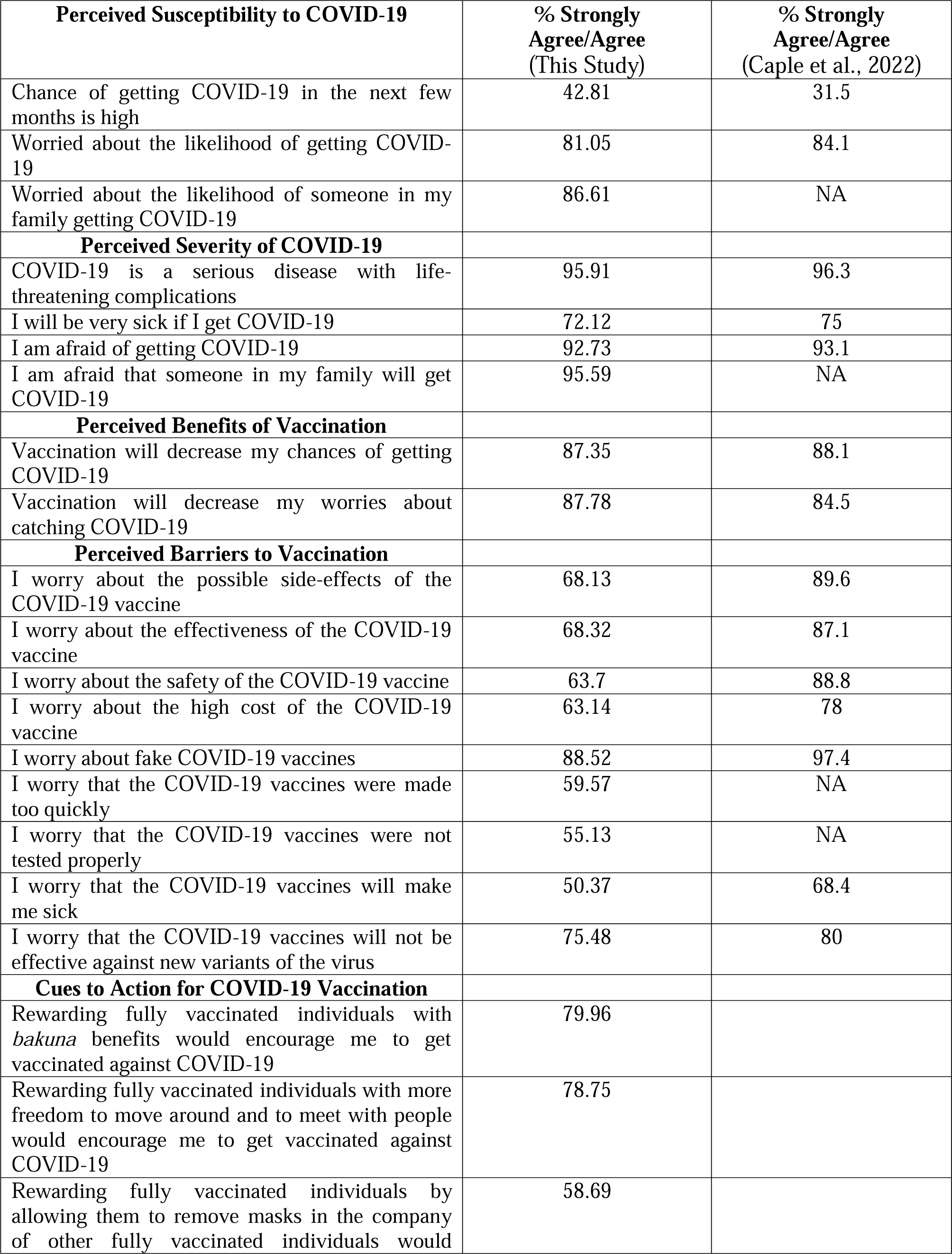
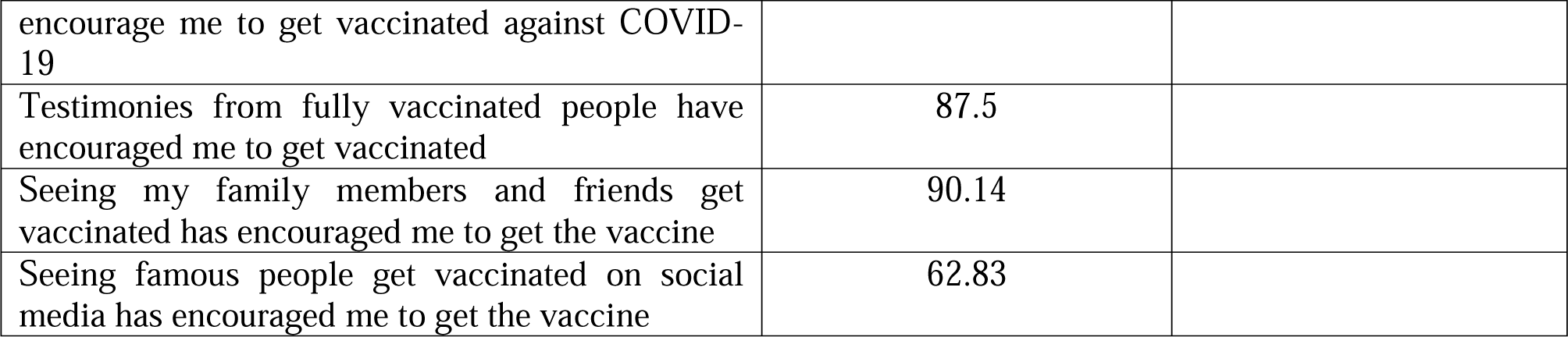
RESPONDENTS’ HEALTH BELIEFS REGARDING COVID-19 AND ITS VACCINES.

With regards to the perceived benefits of vaccination, a majority of participants reported vaccination would decrease their chances of personally contracting COVID-19 (87.35%) as well as their worries about contracting COVID-19 (87.78%). While a majority of respondents reported they had worries about possible side-effects (68.1%), effectiveness (68.3%), safety (63.7%), and high cost (63.1%) of a COVID-19 vaccine, these numbers are overall lower compared to previous findings (Caple et al., 2022). Additionally, a majority of participants reported being worried about fake COVID-19 vaccines (88.5%).

Next, we assessed possible cues to action for COVID-19 vaccination in the course of a national vaccination campaign (Table 2). Our results suggest respondents’ motivation to be immunized was predicted by whether they would be rewarded with perks offered by the private sector including discounts at restaurants and other service industries (80.0% agreed to some extent that they were motivated by this incentive), and rewarded with more freedom to move around and meet other people (78.8% agreed to some extent that they were motivated by this incentive). They were less enthusiastic about rewards that gave them freedom to remove masks in the company of fellow vaccinated individuals (58.69% *strongly agree/agree* that they were motivated by this incentive). Respondents also revealed that hearing from fully vaccinated people (87.5% agreed to some extent) and seeing family members and friends get vaccinated (87.5% agreed to some extent) were predictive factors that would encourage one to seek vaccination.

Finally, as shown in Table 3, respondents were also prompted with statements about COVID-19 perceptions compared to six months prior. A majority of participants agreed to some extent (73%) that their chance of personally contracting COVID-19 was lower compared to six months prior. Similarly, 67.2% of participants agreed to some extent that they were less worried about the likelihood of contracting COVID-19. A majority (63.2%) also revealed that were also less worried about the likelihood of someone in their family contracting COVID-19 compared to six months prior.

**TABLE 3:** CHANGES **IN RESPONDENTS’ HEALTH BELIEFS REGARDING COVID-19 AND ITS VACCINES**

| **Perceived Susceptibility to COVID-19** | **% Strongly Agree/Agree** |
| --- | --- |
| Chance of getting COVID-19 now is lower than it was six months ago | 73 |
| Less worried about the likelihood of getting COVID-19 now than I was six months ago | 67.2 |
| Less worried about the likelihood of someone in my family getting COVID-19 now than I was six months ago | 63.2 |
| Less afraid of getting COVID-19 now than I was six months ago | 63.8 |
| Less afraid that someone in my family will get COVID-19 now than I was six months ago | 61.2 |
| **Perceived Barriers to Vaccination** |  |
| I worry less about the possible side-effects of the COVID-19 vaccine now than I was six months ago | 78.8 |
| I worry less about the effectiveness of the COVID-19 vaccine now than I was six months ago | 76.7 |
| I worry less about the safety of the COVID-19 vaccine now than I was six months ago | 78.6 |
| I worry less about fake COVID-19 vaccines now than I was six months ago | 64.4 |
| I worry less that the COVID-19 vaccines were not tested properly now than I was six months ago | 70.6 |
| I worry less that the COVID-19 vaccines were made too quickly now than I was six months ago | 70.2 |
| I worry less that the COVID-19 vaccines will make me sick now than I was six months ago | 72.0 |
| I worry less that the COVID-19 vaccines will not be effective against new variants of the virus now than I was six months ago | 62.2 |
| **Vaccination Intent** | **%Definitely Yes/Probably Yes** |
| Are you more willing to take a COVID-19 vaccine now than you were 6 months ago? | 92.3 |

With regards to perceived barriers to vaccination, majorities of respondents revealed that they were less worried about the possible side-effects of the vaccines (78.8%), less worried about vaccine effectiveness (76.7%), less worried about vaccine safety (78.64%), less worried about fake vaccines (64.4%), and less worried about vaccine testing (70.57%) compared to six months prior. Overall, our respondents were less worried about every aspect of possible concern associated with COVID-19 vaccination.

### COVID-19 Vaccination Intent

Survey participants were asked whether they had been vaccinated against COVID-19, which was recoded into three groups: not vaccinated (16.7%), partially vaccinated (28.8%), or fully vaccinated (54.5%) (Figure 2A). We also included a specific question to interrogate vaccination intent. As shown in Figure 2B, when responses to the question, “Would you take a COVID-19 vaccine?”, were recoded into two groups, a majority of respondents (95.3%) indicated vaccine acceptance (responses included ‘probably yes’ and ‘definitely yes’). Only 242 of the 5,130 respondents (4.7%) were vaccine resistant (responses included were ‘definitely no’, ‘probably no’, and ‘unsure’). A similar pattern emerged when participants were asked if they were more willing to take a COVID-19 vaccine now as compared to six months prior, with a vast majority (92.3%) identifying as open (responses included ‘definitely yes’ or ‘probably yes’) to the COVID-19 vaccines (Table 3).

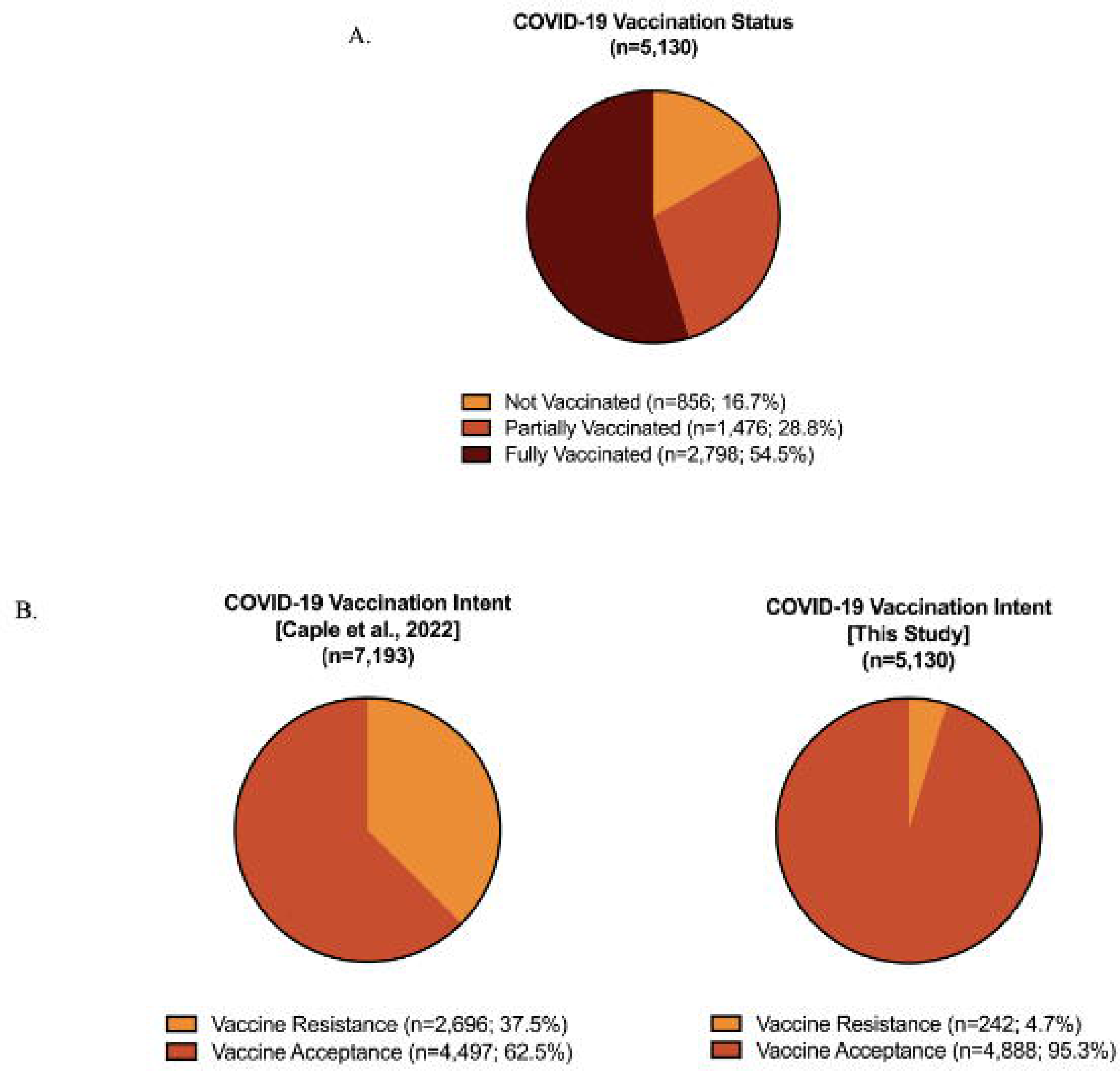

Table 4 shows the univariate and multinomial analyses of factors associated with vaccination status by demographics and HBM constructs. Univariate analyses were conducted examining vaccine resistance and vaccine acceptance as a binary––not vaccinated and partially/fully vaccinated respectively. A total of 4,274 of respondents (83.3%) reported they were either partially (*n* = 1476, 28.8%) or fully vaccinated (*n* = 2798, 54.5%); demonstrating they were vaccine acceptant––while 856 of respondents (16.7%) reported they were not vaccinated suggesting that they were vaccine resistant. The majority of respondents who identified as being in the 18-30 age range, also reported being either partially or fully vaccinated (*n* = 2752, 79.7%).

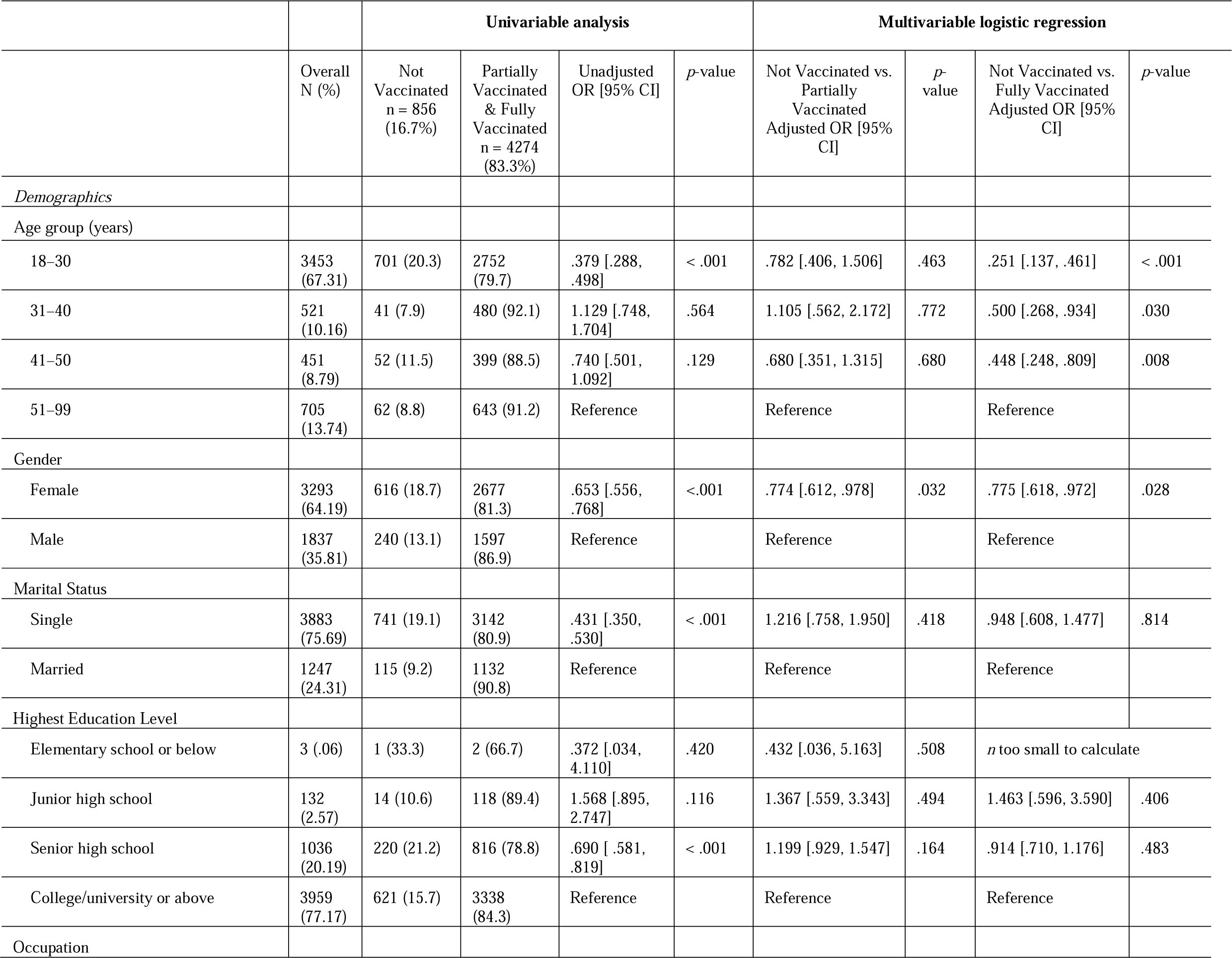
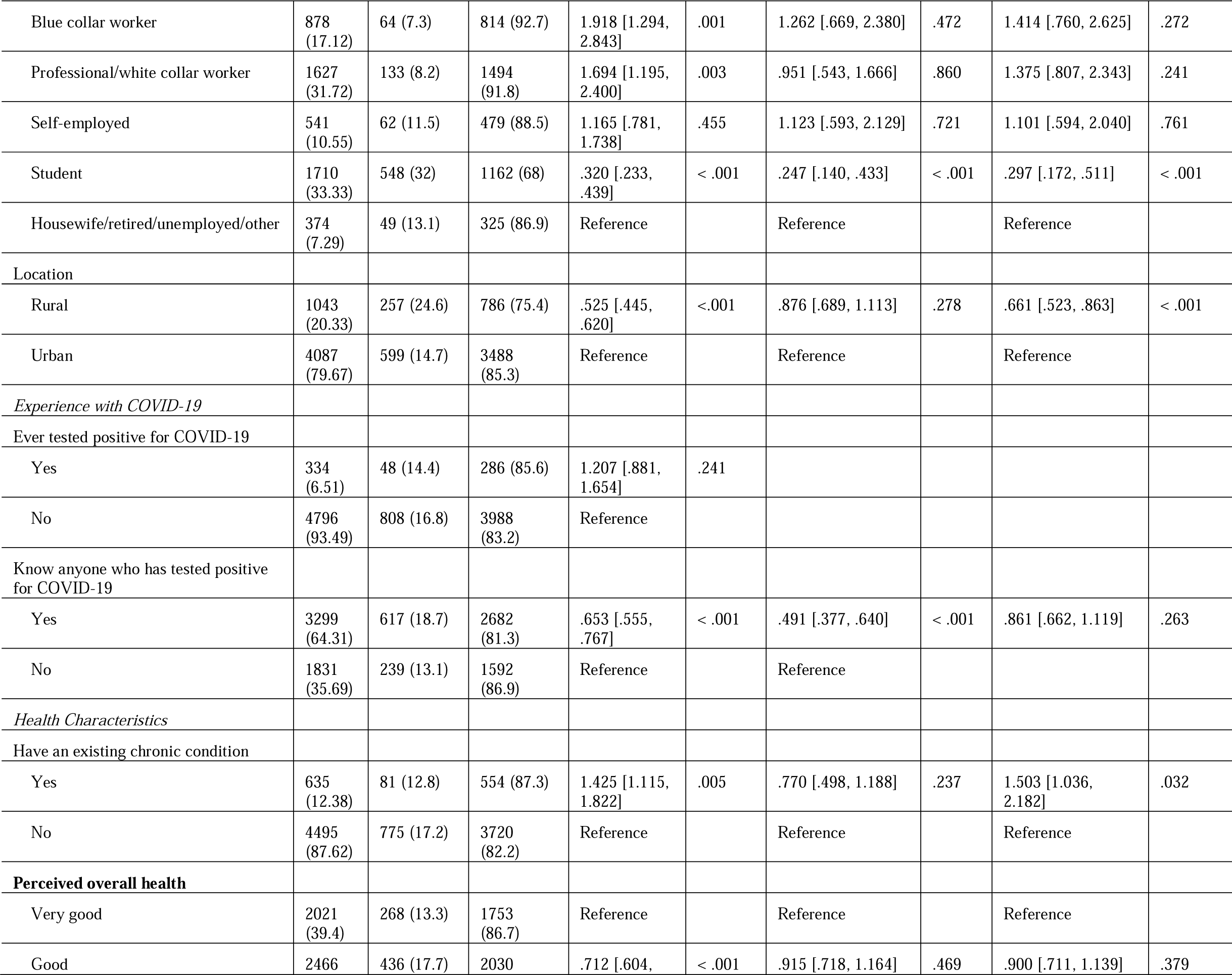
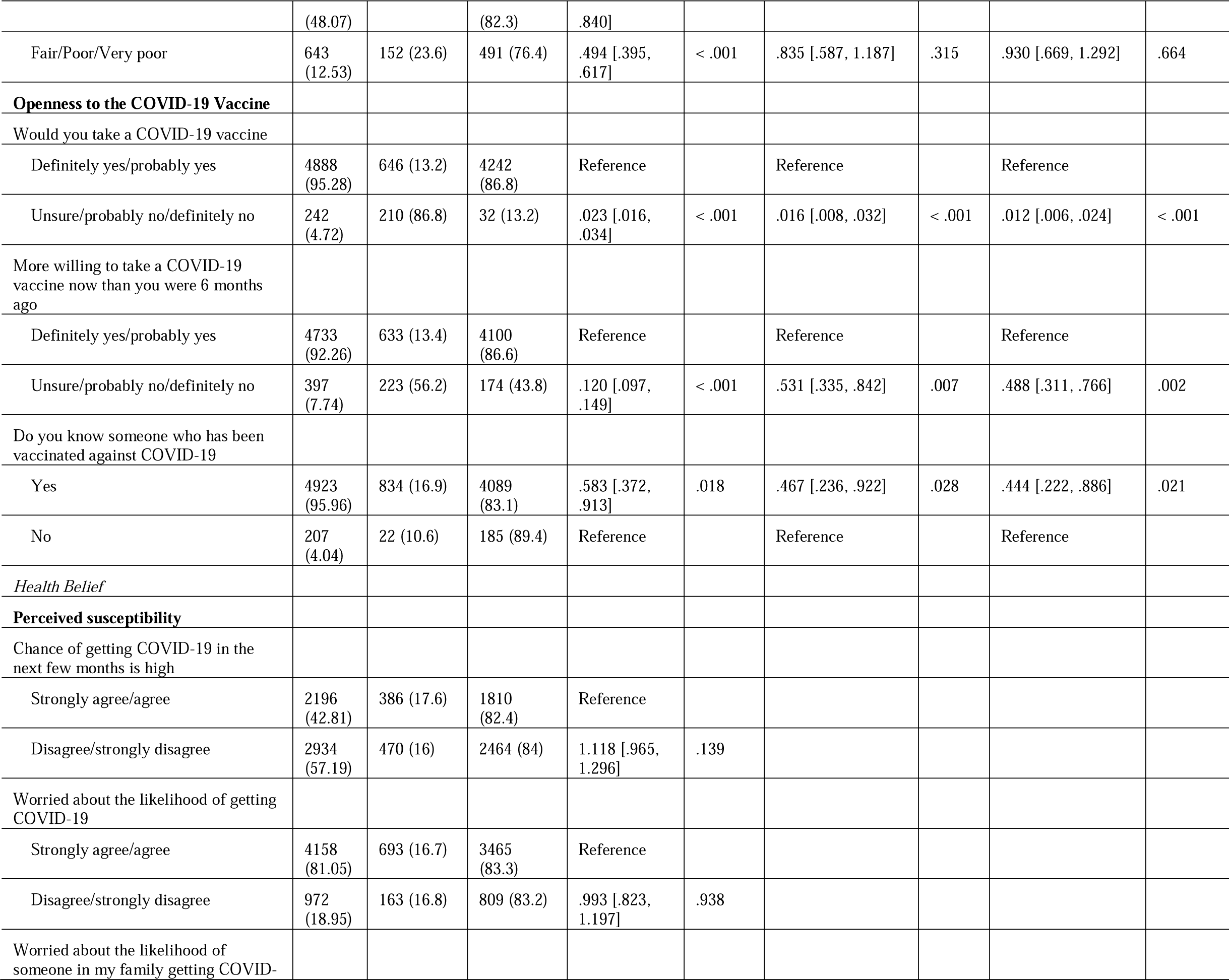
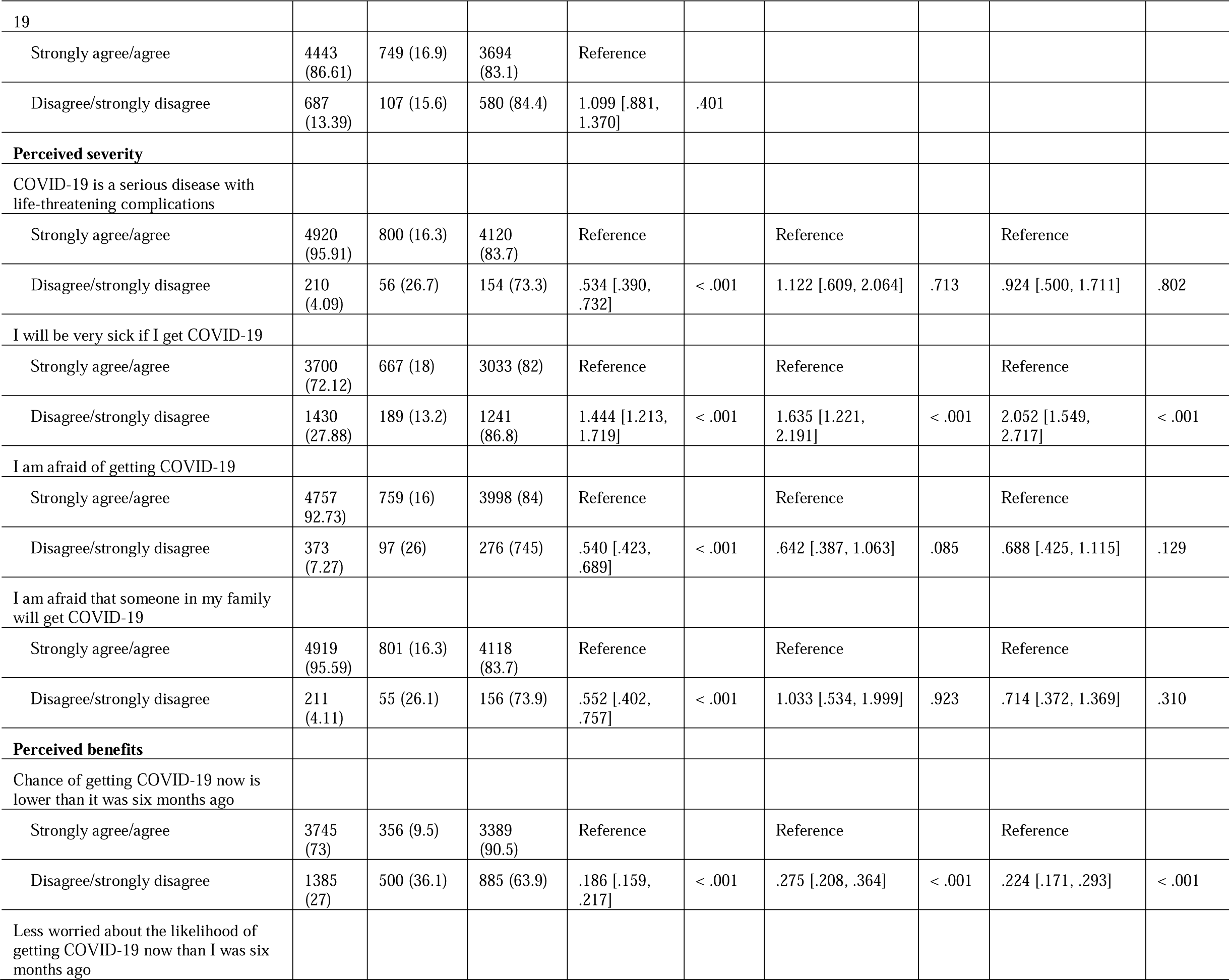
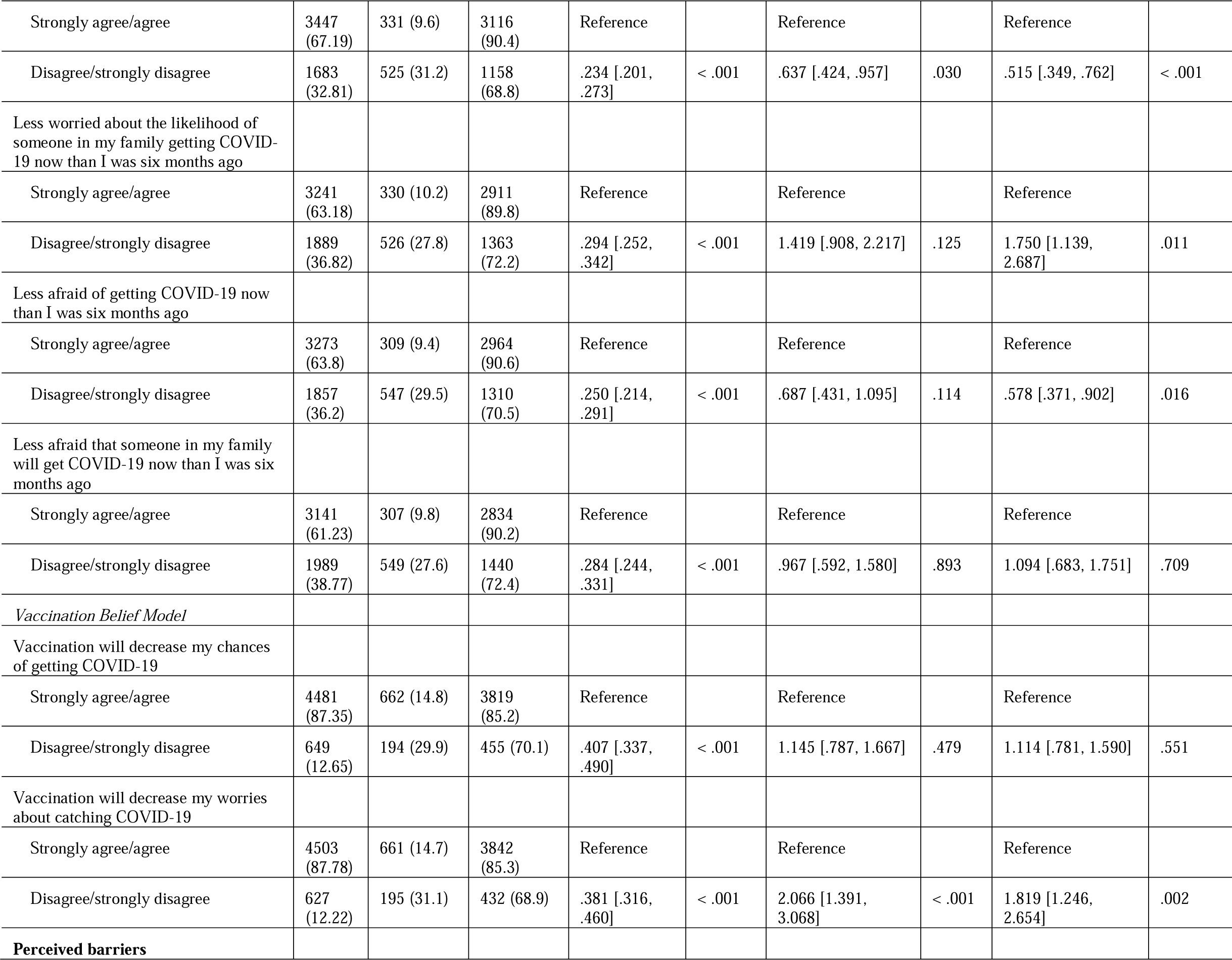
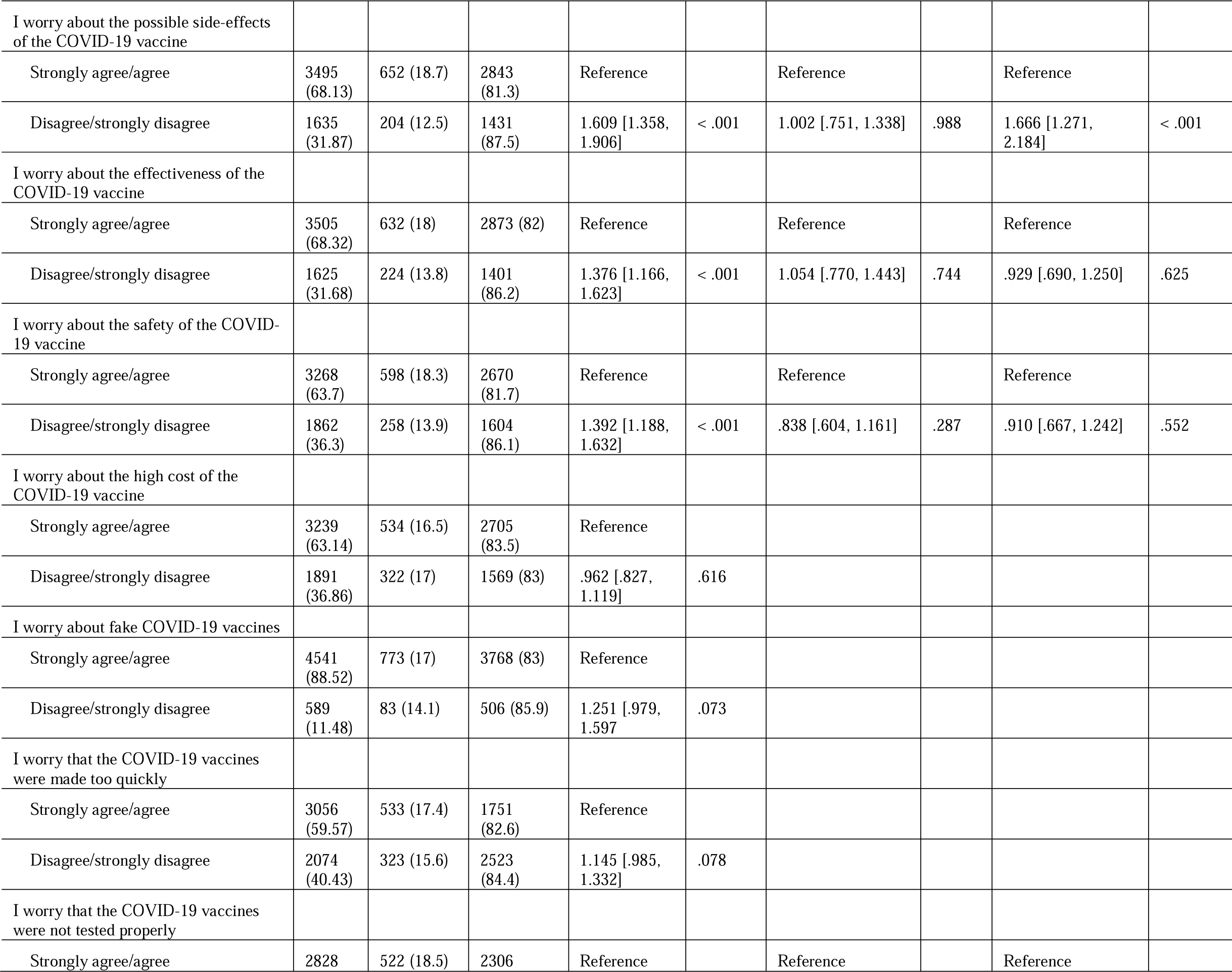
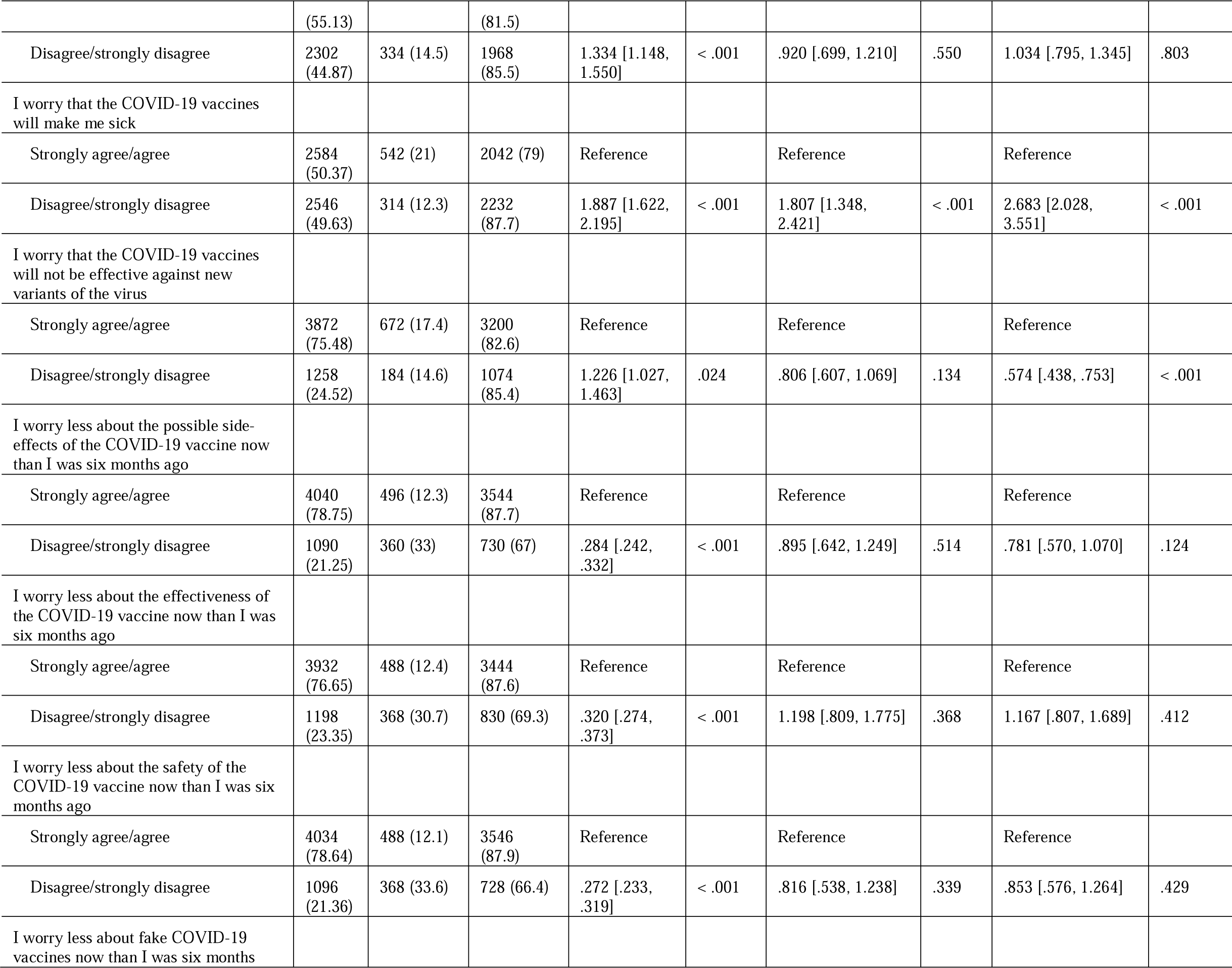
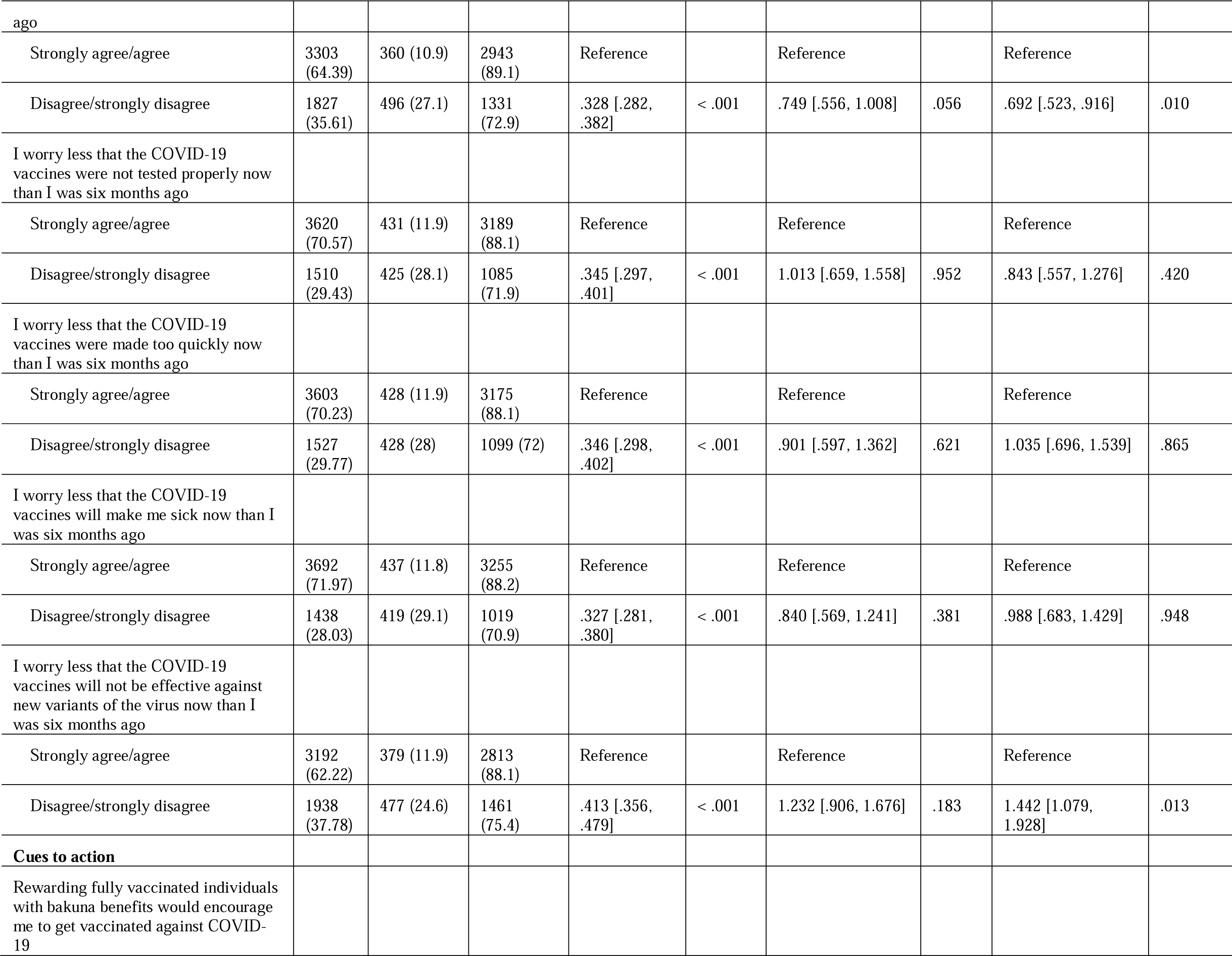
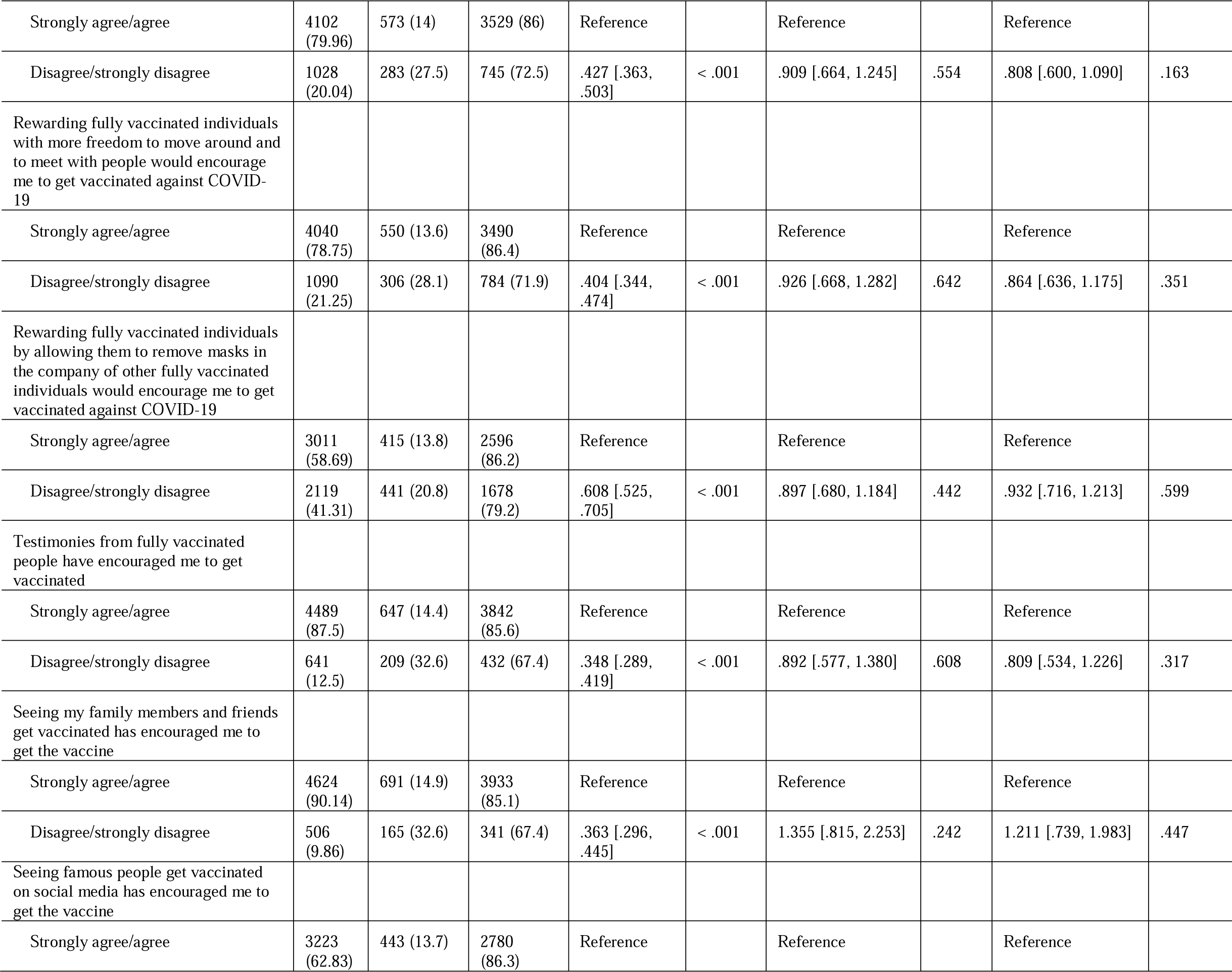
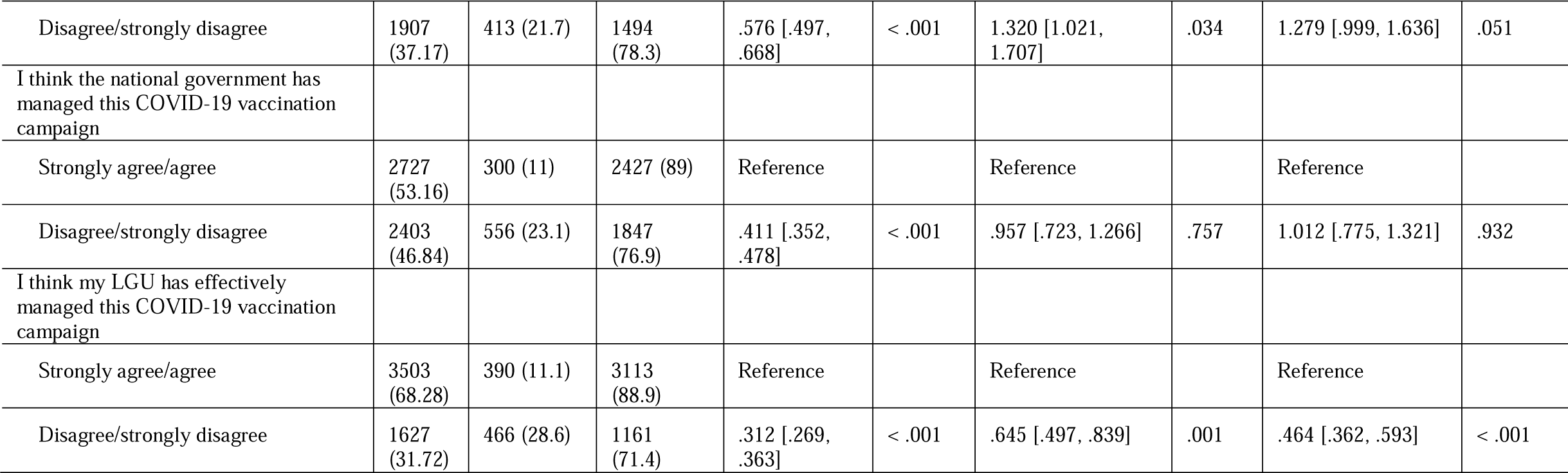

Univariate analyses showed a high proportion of participants who identified as married (90.8%) noted they were either partially or fully vaccinated than compared to 80.9% of partially or fully vaccinated, single respondents. This association, however, was not significant in the multinomial analysis. When examining participant occupation, a higher proportion of respondents that identified as having vaccine acceptance (i.e., either partially or fully vaccinated) also identified as professional/white collar workers (91.2%). Additionally, significant differences were noted in vaccination status by geographic location, where individuals in urban locations (85.3%) reported being either partially or fully vaccinated, while 75.4% of participants in rural locations noted they were either partially or fully vaccination.

Based on demographics, univariate analyses revealed that females have greater odds of being either partially or fully vaccinated (OR = .653, 95% CI [.556, .768]) than males––a finding that also was statistically significant in the multinomial analysis. Furthermore, being a blue collar worker (OR = 1.918, 95% CI [1.294, 2.843]), a professional or white collar worker (OR = 1.694, 95% CI [1.195, 2.400]), a student (OR = .320, 95% CI [.233, .439]), having an existing chronic condition (OR =1.425, 95% CI [1.115, 1.822]), and perceived overall health reported as ‘fair’, ‘poor’, or ‘very poor’ (OR = .494, 95% CI [.395, .617]) and as ‘good’ (OR = .712, 95% CI [.604, .840]), were significant predictors of vaccination status.

Most of the HBM factors were significantly associated with vaccination status. We observed a similar pattern in our previous study (Caple et al., 2022). While 50.4% of respondents noted they were worried COVID-19 vaccines would make them sick, disagreement with that notion (OR = 2.683, 95% CI [2.028, 3.551]) was the strongest predictor of vaccination status when comparing not vaccinated participants to fully vaccinated participants. Individual disagreement with the belief that COVID-19 vaccination would decrease participant worries about personally contracting COVID-19 for not vaccinated compared to partially vaccinated respondents (OR = 2.066, 95% CI [1.391, 3.068]) and for not vaccinated compared to fully vaccinated respondents (OR = 1.819, 95% CI [1.246, 2.654]) was also a strong predictor of vaccination status.

Finally, in Table 5, we compared the HBM constructs that were associated with vaccination intent in our earlier study (Caple et al., 2022), with the same constructs in this study, to determine if they had changed. Our analysis revealed that beliefs around the perceived susceptibility to getting COVID-19--whether the respondent believed that he or she would become sick in the next six months--which were predictive of vaccination intent in our earlier study, were now not able to forecast a respondent’s openness to the vaccines. Strikingly, two more belief constructs regarding the perceived barriers of expensive vaccines and of fake vaccines, also ceased being predictive in this study when they were predictive in our earlier report.

|  |  | Factors associated with COVID-19 vaccine status after national vaccine rollout |  |  | Factors Associated with intention to take the COVID-19 vaccine before national vaccine rollout (Caple et al., 2022) |  |  |  |
| --- | --- | --- | --- | --- | --- | --- | --- | --- |
|  | Overall N (%) | Not Vaccinated<br>n = 856<br>(16.7%) | Partially Vaccinated & Fully Vaccinated<br>n = 4274<br>(83.3%) | Individual Predictor of vaccine status? | Overall N (%) | Vaccine hesitant<br>n = 2696 (%) | Not vaccine hesitant<br>n = 4497 (%) | Individual Predictor of vaccine intent? |
| *Demographics* |  |  |  |  |  |  |  |  |
| Age group (years) |  |  |  | Yes |  |  |  | No |
| 18–30 | 3453<br>(67.31) | 701 (20.3) | 2752 (79.7) |  | 3770<br>(52.4) | 1405 (37.3) | 2365<br>(62.7) |  |
| 31–40 | 521<br>(10.16) | 41 (7.9) | 480 (92.1) |  | 815<br>(11.3) | 306 (37.5) | 509 (62.5) |  |
| 41–50 | 451<br>(8.79) | 52 (11.5) | 399 (88.5) |  | 861<br>(12) | 335 (38.9) | 526 (61.1) |  |
| 51–99 | 705<br>(13.74) | 62 (8.8) | 643 (91.2) |  | 1747<br>(24.29) | 650 (37.21) | 1097<br>(62.79) |  |
| Gender |  |  |  | Yes |  |  |  | Yes |
| Female | 3293<br>(64.19) | 616 (18.7) | 2677 (81.3) |  | 4789<br>(66.6) | 1955 (40.8) | 2834<br>(59.2) |  |
| Male | 1837<br>(35.81) | 240 (13.1) | 1597 (86.9) |  | 2404<br>(33.4) | 741 (30.8) | 1663<br>(69.2) |  |
| Marital Status |  |  |  | Yes |  |  |  | Yes |
| Single | 3883<br>(75.69) | 741 (19.1) | 3142 (80.9) |  | 4724<br>(65.7) | 1719 (36.4) | 3005<br>(63.6) |  |
| Married | 1247<br>(24.31) | 115 (9.2) | 1132 (90.8) |  | 2469<br>(34.3) | 977 (39.6) | 1492<br>(60.4) |  |
| Highest Education Level |  |  |  | Yes |  |  |  | Yes |
| Elementary school or below | 3 (.06) | 1 (33.3) | 2 (66.7) |  | 1 | 1 |  |  |
| Junior high school | 132<br>(2.57) | 14 (10.6) | 118 (89.4) |  | 46 (.6) | 29 (63) | 17 (37) |  |
| Senior high school | 1036<br>(20.19) | 220 (21.2) | 816 (78.8) |  | 1102<br>(15.3) | 378 (34.3) | 724 (65.7) |  |
| College/university or above | 3959<br>(77.17) | 621 (15.7) | 3338 (84.3) |  | 6044<br>(84) | 2288 (37.9) | 3756<br>(62.1) |  |
| Occupation |  |  |  | Yes |  |  |  | Yes |
| Blue collar worker | 878<br>(17.12) | 64 (7.3) | 814 (92.7) |  | 91<br>(1.3) | 42 (46.2) | 49 (53.8) |  |
| Professional/white collar worker | 1627<br>(31.72) | 133 (8.2) | 1494 (91.8) |  | 2980<br>(41.4) | 1133 (38) | 1847 (62) |  |
| Self-employed | 541<br>(10.55) | 62 (11.5) | 479 (88.5) |  | 575 (8) | 254 (44.2) | 321 (55.8) |  |
| Student | 1710<br>(33.33) | 548 (32) | 1162 (68) |  | 2831<br>(39.4) | 976 (34.5) | 1855<br>(65.5) |  |
| Housewife/retired/unemployed/other | 374<br>(7.29) | 49 (13.1) | 325 (86.9) |  | 716<br>(10) | 291 (40.6) | 425 (59.4) |  |
| Location |  |  |  | Yes |  |  |  | Yes |
| Rural | 1043<br>(20.33) | 257 (24.6) | 786 (75.4) |  | 1517<br>(21.1) | 711 (46.9) | 806 (53.1) |  |
| Urban | 4087 | 599 (14.7) | 3488 (85.3) |  | 5676 | 1985 (35) | 3691 (65) |  |
|  | (79.67) |  |  |  | (78.9) |  |  |  |
| *Experience with COVID-19* |  |  |  |  |  |  |  |  |
| Ever tested positive for COVID-19 |  |  |  | No |  |  |  | No |
| Yes | 334<br>(6.51) | 48 (14.4) | 286 (85.6) |  | 211<br>(2.9) | 68 (32.2) | 143 (67.8) |  |
| No | 4796<br>(93.49) | 808 (16.8) | 3988 (83.2) |  | 6982<br>(97.1) | 2628 (37.6) | 4354<br>(62.4) |  |
| Know anyone who has tested positive for COVID-19 |  |  |  | Yes |  |  |  | Yes |
| Yes | 3299<br>(64.31) | 617 (18.7) | 2682 (81.3) |  | 5234<br>(72.8) | 1780 (34) | 3454 (66) |  |
| No | 1831<br>(35.69) | 239 (13.1) | 1592 (86.9) |  | 1959<br>(27.2) | 916 (46.8) | 1043<br>(53.2) |  |
| *Health Characteristics* |  |  |  |  |  |  |  |  |
| Have an existing chronic condition |  |  |  | Yes |  |  |  | Yes |
| Yes | 635<br>(12.38) | 81 (12.8) | 554 (87.3) |  | 1178<br>(16.4) | 396 (33.6) | 782 (66.4) |  |
| No | 4495<br>(87.62) | 775 (17.2) | 3720 (82.2) |  | 6015<br>(83.6) | 2300 (38.2) | 3715<br>(61.8) |  |
| **Perceived overall health** |  |  |  | Yes |  |  |  | Yes |
| Very good | 2021<br>(39.4) | 268 (13.3) | 1753 (86.7) |  | 1997<br>(27.8) | 802 (40.2) | 1195<br>(59.8) |  |
| Good | 2466<br>(48.07) | 436 (17.7) | 2030 (82.3) |  | 3934<br>(54.7) | 1376 (35) | 2558 (65) |  |
| Fair/Poor/Very poor | 643<br>(12.53) | 152 (23.6) | 491 (76.4) |  | 1262<br>(17.5) | 518 (41) | 744 (59) |  |
| *Health Belief* |  |  |  |  |  |  |  |  |
| **Perceived susceptibility** |  |  |  |  |  |  |  |  |
| Chance of getting COVID-19 in the next few months is high |  |  |  | No |  |  |  | Yes |
| Strongly agree/agree | 2196<br>(42.81) | 386 (17.6) | 1810 (82.4) |  | 2264<br>(31.5) | 1595 (70.5) | 669 (29.5) |  |
| Disagree/strongly disagree | 2934<br>(57.19) | 470 (16) | 2464 (84) |  | 4929<br>(68.5) | 2027 (41.1) | 2902<br>(58.9) |  |
| Worried about the likelihood of getting COVID-19 |  |  |  | No |  |  |  | Yes |
| Strongly agree/agree | 4158<br>(81.05) | 693 (16.7) | 3465 (83.3) |  | 6047<br>(84.1) | 2105 (34.8) | 3942<br>(65.2) |  |
| Disagree/strongly disagree | 972<br>(18.95) | 163 (16.8) | 809 (83.2) |  | 1146<br>(15.9) | 591 (51.6) | 555 (48.4) |  |
| **Perceived severity** |  |  |  |  |  |  |  |  |
| COVID-19 is a serious disease with life-threatening complications |  |  |  | Yes |  |  |  | Yes |
| Strongly agree/agree | 4920<br>(95.91) | 800 (16.3) | 4120 (83.7) |  | 6928<br>(96.32) | 2523 (36.4) | 4405<br>(63.6) |  |
| Disagree/strongly disagree | 210<br>(4.09) | 56 (26.7) | 154 (73.3) |  | 265<br>(3.68) | 173 (65.3) | 95 (34.7) |  |
| I will be very sick if I get COVID-19 |  |  |  | Yes |  |  |  | Yes |
| Strongly agree/agree | 3700<br>(72.12) | 667 (18) | 3033 (82) |  | 5394<br>(75) | 1911 (35.4) | 3483<br>(64.6) |  |
| Disagree/strongly disagree | 1430 | 189 (13.2) | 1241 (86.8) |  | 1799 | 785 (43.6) | 1014 |  |
|  | (27.88) |  |  |  | (25) |  | (56.4) |  |
| I am afraid of getting COVID-19 |  |  |  | Yes |  |  |  | Yes |
| Strongly agree/agree | 4757<br>(92.73) | 759 (16) | 3998 (84) |  | 6700<br>(93.15) | 2439 (36.4) | 4261<br>(63.6) |  |
| Disagree/strongly disagree | 373<br>(7.27) | 97 (26) | 276 (745) |  | 493<br>(6.85) | 257 (52.1) | 236 (47.9) |  |
| *Vaccination Belief Model* |  |  |  |  |  |  |  |  |
| Vaccination will decrease my chances of getting COVID-19 |  |  |  | Yes |  |  |  | Yes |
| Strongly agree/agree | 4481<br>(87.35) | 662 (14.8) | 3819 (85.2) |  | 6339<br>(88.1) | 2006 (31.6) | 4333<br>(68.4) |  |
| Disagree/strongly disagree | 649<br>(12.65) | 194 (29.9) | 455 (70.1) |  | 854<br>(11.9) | 690 (80.8) | 164 (19.2) |  |
| Vaccination will decrease my worries about catching COVID-19 |  |  |  | Yes |  |  |  | Yes |
| Strongly agree/agree | 4503<br>(87.78) | 661 (14.7) | 3842 (85.3) |  | 6078<br>(84.5) | 1833 (30.2) | 4245<br>(69.8) |  |
| Disagree/strongly disagree | 627<br>(12.22) | 195 (31.1) | 432 (68.9) |  | 1115<br>(15.5) | 863 (77.4) | 252 (22.6) |  |
| **Perceived barriers** |  |  |  |  |  |  |  |  |
| I worry about the possible side-effects of the COVID-19 vaccine |  |  |  | Yes |  |  |  | Yes |
| Strongly agree/agree | 3495<br>(68.13) | 652 (18.7) | 2843 (81.3) |  | 6447<br>(89.6) | 2647 (41.1) | 3800<br>(58.9) |  |
| Disagree/strongly disagree | 1635<br>(31.87) | 204 (12.5) | 1431 (87.5) |  | 746<br>(10.4) | 49 (6.6) | 697 (93.4) |  |
| I worry about the effectiveness of the COVID-19 vaccine |  |  |  | Yes |  |  |  | Yes |
| Strongly agree/agree | 3505<br>(68.32) | 632 (18) | 2873 (82) |  | 6263<br>(87.1) | 2581 (41.2) | 3682<br>(58.8) |  |
| Disagree/strongly disagree | 1625<br>(31.68) | 224 (13.8) | 1401 (86.2) |  | 930<br>(12.9) | 115 (12.4) | 815 (87.6) |  |
| I worry about the safety of the COVID-19 vaccine |  |  |  | Yes |  |  |  | Yes |
| Strongly agree/agree | 3268<br>(63.7) | 598 (18.3) | 2670 (81.7) |  | 6389<br>(88.8) | 2626 (41.1) | 3763<br>(58.9) |  |
| Disagree/strongly disagree | 1862<br>(36.3) | 258 (13.9) | 1604 (86.1) |  | 804<br>(11.2) | 70 (8.7) | 734 (91.3) |  |
| I worry about the high cost of the COVID-19 vaccine |  |  |  | No |  |  |  | Yes |
| Strongly agree/agree | 3239<br>(63.14) | 534 (16.5) | 2705 (83.5) |  | 5608<br>(78) | 2301 (41) | 3307 (59) |  |
| Disagree/strongly disagree | 1891<br>(36.86) | 322 (17) | 1569 (83) |  | 1585<br>(22) | 395 (24.9) | 1190<br>(75.1) |  |
| I worry about fake COVID-19 vaccines |  |  |  | No |  |  |  | Yes |
| Strongly agree/agree | 4541<br>(88.52) | 773 (17) | 3768 (83) |  | 7006<br>(97.4) | 2646 (37.8) | 4360<br>(62.2) |  |
| Disagree/strongly disagree | 589<br>(11.48) | 83 (14.1) | 506 (85.9) |  | 187<br>(.03) | 50 (26.7) | 197 (73.3) |  |
| I worry that the COVID-19 vaccines will make me sick |  |  |  | Yes |  |  |  | Yes |
| Strongly agree/agree | 2584 | 542 (21) | 2042 (79) |  | 4918 | 2404 (48.9) | 2514 |  |
|  | (50.37) |  |  |  | (68.4) |  | (51.1) |  |
| Disagree/strongly disagree | 2546<br>(49.63) | 314 (12.3) | 2232 (87.7) |  | 2275<br>(31.6) | 292 (12.8) | 1983<br>(87.2) |  |
| I worry that the COVID-19 vaccines<br>will not be effective against new<br>variants of the virus |  |  |  | Yes |  |  |  | Yes |
| Strongly agree/agree | 3872<br>(75.48) | 672 (17.4) | 3200 (82.6) |  | 5757<br>(80) | 2506 (43.5) | 3251<br>(56.5) |  |
| Disagree/strongly disagree | 1258<br>(24.52) | 184 (14.6) | 1074 (85.4) |  | 1436<br>(20) | 190 (13.2) | 1246<br>(86.8) |  |

## Discussion

How does an ongoing vaccination campaign influence vaccination intent, especially during a pandemic? How do the presence of vaccines and vaccinated individuals shape and alter, if at all, vaccination intent? In our previous study, completed two months *before* the rollout of the national COVID-19 vaccination campaign in the Philippines, we showed that Health Belief Model (HBM) constructs were significantly associated with vaccination intent in the archipelago (Caple et al., 2022). Perceptions of high susceptibility, high severity, and significant benefits were all good predictors for vaccination intent. We also found that external cues to action were important.

Here, in a follow-up study completed six months *after* the beginning of the national vaccination campaign, we discovered that the presence of vaccines and vaccinated individuals in a Filipino community predicted vaccine intent. Most significantly, in our earlier study, 62.5% of our respondents indicated that they were willing to be vaccinated when the vaccines became available (Caple et al., 2022). This study revealed that at the time of our survey, 83.3% of our respondents had already begun the vaccination series (Figure 2). Strikingly, even more individuals (95.3%) indicated that they were willing to be vaccinated. When directly asked if they were more willing to receive a vaccination than six months prior, 92.3% agreed to some extent that they were indeed more willing. This significant increase in vaccination intent during a comparable time period has also been observed by others in the Philippines (Macaraan, 2021).

How do we explain the difference in numbers between those persons who were vaccinated (83.3%) and those who said that they were willing to be vaccinated (95.3%)? There are at least two possibilities. First, there could have been a group of individuals who were still waiting for vaccine access: These Filipinos were willing to be vaccinated, but there were no vaccines for them at the time of the survey. This scenario is likely because a significant minority (20.33%) of our respondents lived in rural areas. At the time of our survey, six months into the national vaccination campaign, the Philippines was still prioritizing individuals in the urban areas of the country in a bid to build a wall of immunization around cities that would prevent the transmission of the SARS-CoV2 virus into the rural countryside. This public policy was named the NCR+8 vaccination strategy (Casinas, 2021; Almajose et al., 2021).

Second, there could have been a group of individuals who were waiting to be vaccinated, allowing others to be vaccinated ahead of them, because of the shortage of vaccines. Though there were anecdotal stories of altruistic persons circulating on social media during the earliest months of the national vaccination campaign in the Philippines, they have never been confirmed.

Most notably, our study reveals that the presence of vaccines and vaccinated individuals in a Filipino community predicts vaccination intent (Figure 2). Note that it cannot simply be the presence of vaccines or vaccinated individuals that predicted vaccination intent, because the global vaccination campaign had already begun at the time of our first study, unleashing numerous posts and reports on Filipino social media platforms of vaccinated citizens in the Global North. As our study revealed, what is significant for predicting vaccination intent is not whether or not the respondents knew that others were getting vaccinated. Rather what was most important is that our respondents personally knew loved ones and friends who had been vaccinated (Table 2). These were the most significant cues of action to COVID-19 vaccination in our study. Similarly, a COVID-19 study in Taiwan has also reported the effect of positive word-of-mouth testimony on vaccination intent (Wu & Chiang, 2023).

What is the significance of this finding? It suggests that Filipinos want to hear and to trust the testimonies of their *kababayans* (“fellow Filipinos, townmates, or countrymen”). Thus, future vaccination campaigns in the Philippines should showcase Filipinos who have been vaccinated, especially those who live in the same *barangay* as the target vaccination population. A *barangay* is the smallest administrative division in the Philippines and is the native Filipino term for a village, a district, or ward. The more familiar the testimony of the vaccinated *kababayan*, the better.

As for additional cues to action for COVID-19 vaccination, our study revealed that our Filipino respondents believed that concrete rewards for immunization like the private sector’s *bakuna* benefits program which gave immunized individuals exclusive perks, discounts, and even freebies, would motivate them to get immunized. These *bakuna* benefits included free menu items, free upgrades on drink sizes, and “buy one, get one” promotional items (Ignacio, 2021). Other vaccination perks including more freedom to mingle with others also motivated our respondents (78.8% agreed to some extent that this would encourage them) though incentives that would give them more freedom to remove masks would not motivate them as much (58.69% agreed to some extent that this would encourage them). Why not masks? Surveys have shown that Filipinos value masking and that significant numbers of them intended to continue wearing their masks even years after the official end of the pandemic (De Vera-Ruiz, 2022).

Next, despite the changes in the numbers of respondents who became more open to the COVID-19 vaccines after the rollout of the national vaccination campaign, there were no significant changes in the predictive power of the HBM constructs during the same time period. For the most part, HBM factors that predicted vaccination intent in our earlier study also predicted vaccination intent in this current study (Table 5). Only one, age of the respondent, became predictive when it was not so in our previous work. One simple explanation for this is that the vaccination rollout in the Philippines prioritized the elderly. As such, at the time of our survey, it was significantly more likely for a senior citizen to be vaccinated than for a young adult to have been immunized (Rappler.com, 2021). This was not the case prior to the arrival of the vaccines.

Four of the twenty-four HBM constructs assessed in the current study no longer predicted vaccine intent once the national vaccination strategy was underway (Table 5). Two involved a respondent’s perceived susceptibility to COVID-19, and two were related to one’s worries regarding the cost and the authenticity of the COVID-19 vaccines. In our earlier study, respondents who believed that there was a high risk that they would get infected with SARS-CoV2 were more likely to be open to the COVID-19 vaccines (Caple et al., 2022). In this study, perceived susceptibility ceased to be associated with a positive vaccine intent. One possible explanation is that a large number of our respondents in this study (67.2%) also reported that they had simply become less worried about COVID-19. These results suggest that fear of severe illness simply became less of a concern among our respondents as the vaccines were rolled out.

Similarly, worries surrounding the cost and authenticity of the COVID-19 vaccines lost their ability to predict vaccination intent. This should not be surprising since the availability of free and legitimate vaccines in the Philippines would have made these concerns moot in the minds of many Filipinos. Of note, the national government launched a widespread public advisory campaign against fake COVID-19 vaccines throughout the rollout of the national vaccination campaign. It was quick to announce investigations of companies purporting to distribute fake vaccines (CNN, 2021).

Finally, our study has several limitations. As we already noted with our earlier study, the use of an open-access online survey may result in sampling bias so we cannot generalize our findings to the entire Filipino population (Wyatt, 2000; Eysenbach & Wyatt, 2002). Furthermore, the responses were based on self-report and may be subject to self-reporting bias and a tendency to report socially desirable responses especially in a strongly collectivist society like the Philippines. Despite these shortcomings, we believe that our findings will provide insights to support future vaccine rollouts in the Philippines by helping public health authorities to understand vaccine demand and vaccine hesitancy in the country.

## Data Availability

All data produced in the present study are available upon reasonable request to the authors. They will be included in the final publication in a peer-reviewed journal.

## Notes

### Competing Interest Statement

The authors have declared no competing interest.

